# Impact of preexisting dementia on in-hospital outcomes in patients with intracerebral hemorrhage stroke in China

**DOI:** 10.1101/2023.08.21.23294393

**Authors:** Lijun Zuo, Yang Hu, YanHong Dong, Hongqiu Gu, Raymond CS Seet, Zixiao Li, Yongjun Wang, Xingquan Zhao

## Abstract

**Objective:** We assessed the impact of preexisting dementia on in-hospital mortality, home discharge and complications of Chinese patients with intracerebral hemorrhage (ICH).

**Methods:** Consecutive in-hospital data were extracted from the China Stroke Center Alliance database from August 2015 to July 2019. Patient characteristics, in-hospital mortality, and complications were compared between ICH patients with and without preexisting dementia.

**Results:** Out of the eligible 72,318 patients with ICH, we identified 328 patients with pre-existing dementia. Compared with patients without dementia, those in the dementia group were older, more females and a higher proportion of individuals with lower education, and a history of diabetes, myocardial infarction, stroke, heart failure, peripheral vascular disease and cigarette smoking. Those with pre-existing dementia group were more prone to a greater stroke severity as measured by the National Institute of Health Stroke Scale (NIHSS) and Glasgow Coma Scale (GCS) at presentation. In the adjusted models, the presence of preexisting dementia was associated with an increased risk of in-hospital mortality (OR 2.31, 95% CI 1.12-4.77) and more frequent in-hospital complications of pulmonary embolism (OR 5.41, 95% CI 1.16-25.14), pneumonia (OR 1.58, 95% CI 1.08-2.33), urinary tract infection (OR 2.37; 95% CI 1.21-4.64), and gastrointestinal bleeding (OR 2.39, 95% CI 1.27-4.49).

**Conclusions:** ICH patients with pre-existing dementia are more prone to more severe strokes and poorer outcomes. Future studies should evaluate the value of intensive risk factor control among individuals with pre-existing dementia for stroke prevention.

## Introduction

Intracerebral hemorrhage (ICH) accounts for 10% to 15% of strokes worldwide and is associated with increased morbidity and mortality^1^. Although pre-existing cognitive impairment has been observed as an important predictor of mortality in stroke patients, previous investigations have mostly focused on patients with ischemic stroke and in Caucasian populations^2-4^. Limited data are available among individuals with ICH and the impact of pre-existing dementia on ICH outcomes is poorly studied in non-Caucasian populations. We examined the impact of pre-existing dementia on ICH outcomes in a Chinese population by leveraging on registry data from the China Stroke Center Alliance.

## Methods and Materials

The China Stroke Center Alliance (CSCA) is a national registry that systematically collects data from 1476 hospitals to identify epidemiological trends of stroke and stroke outcomes in China, where stroke remains a leading cause of death^5^. Between August 2015 to July 2019, data from patients with ICH were included, and those with cerebral venous sinus thrombosis, subarachnoid hemorrhage, or ischemic stroke, were excluded. Information on patient demographics (age and sex), household income, education level and medical history was collected, and stroke severity was assessed using the Glasgow Coma Scale (GCS) and National Institutes of Health Stroke Scale (NIHSS). The primary study endpoint was in-hospital mortality, whilst the secondary endpoints were in-hospital complications such as pneumonia, decubitus ulcer, pulmonary embolism, myocardial infarction, gastrointestinal bleeding, deep vein thrombosis (DVT) and stroke recurrence. The study protocol was approved by the ethics committee of the Beijing Tiantan Hospital (No. KY2015-001-01) and the study was performed in accordance with the Declaration of Helsinki. All participants or their legal representatives provided written informed consent. Continuous and categorical covariates were presented as either mean (standard deviation) or median (interquartile range) values, and in percentages, respectively. Pearson’s χ ^2^ test or Fisher’s exact test was used to determine the group differences for categorical variables. Odds ratios with 95% confidence intervals were derived using logistic regression methods and adjustments for performed for potential confounders. Statistical significance was considered when P<0.05. The SAS 9.4 software (SAS Institute Inc, Cary, NC) was used for all analysis.

## Results

Out of 1,006,798 patients included in the CSCA registry, 814,139 patients who had ischemic stroke/transient ischemic attack, and 8,766 patients with subarachnoid hemorrhage, were excluded. A further 11,582 patients were excluded as their stroke subtype could not be verified and 244 patients whose clinical outcomes were not available were excluded. Data from 72,318 patients formed the study cohort (Figure 1). Their baseline characteristics are summarized in Table 1. Compared with non-dementia controls, ICH patients with pre-existing dementia were older and comprised more females. A higher proportion of patients with pre-existing dementia had lower education, and a history of diabetes, myocardial infarction, stroke, heart failure, peripheral vascular disease, and cigarette smoking. Those with pre-existing dementia group were more prone to a greater stroke severity as measured by the National Institute of Health Stroke Scale (NIHSS) and Glasgow Coma Scale (GCS) at presentation. ICH patients with preexisting dementia had higher levels of creatinine and blood urea nitrogen compared with non-dementia controls.

**Fig.1.**
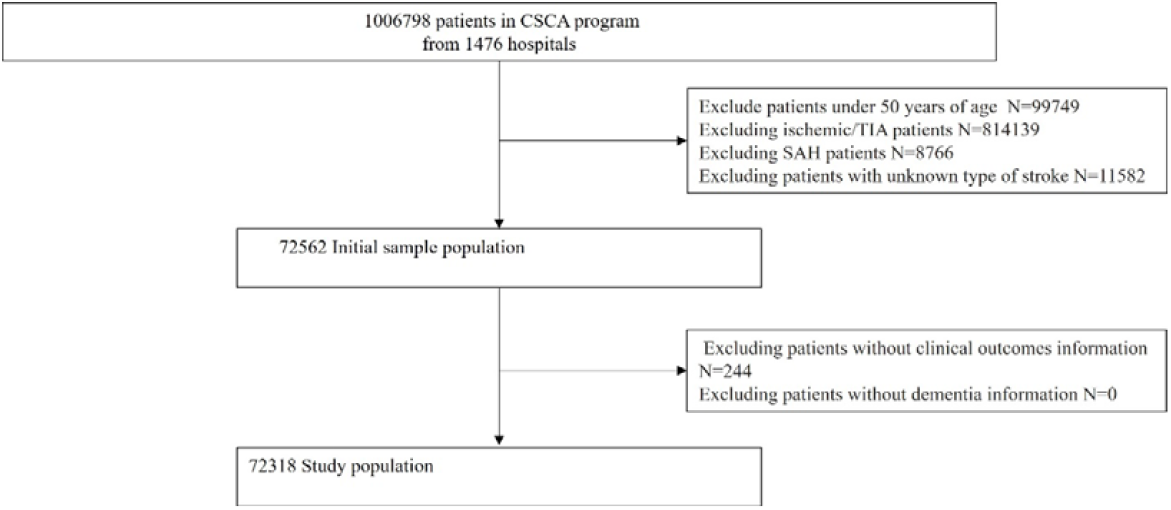
the flowchart of patients participating in the study

In-hospital mortality in ICH patients with pre-existing dementia was significantly higher than that in non-dementia patients. Patients with pre-existing dementia were more prone to in-hospital complications such as pulmonary embolism, pneumonia, urinary tract infection, gastrointestinal bleeding, and hydrocephalus. Patients with preexisting dementia harbor increased risks of recurrent intracerebral hemorrhage (Table 2). In the adjusted models, patients with preexisting dementia have with higher risk of in-hospital mortality (OR 2.31, 95% CI 1.12-4.77) and more frequent in-hospital complications of pulmonary embolism (OR 5.41, 95% CI 1.16-25.14), pneumonia (OR 1.58, 95% CI 1.08-2.33), urinary tract infection (OR 2.37; 95% CI 1.21-4.64), and gastrointestinal bleeding (OR 2.39, 95% CI 1.27-4.49) (Table 3 and Figure 2).

**Fig.2.**
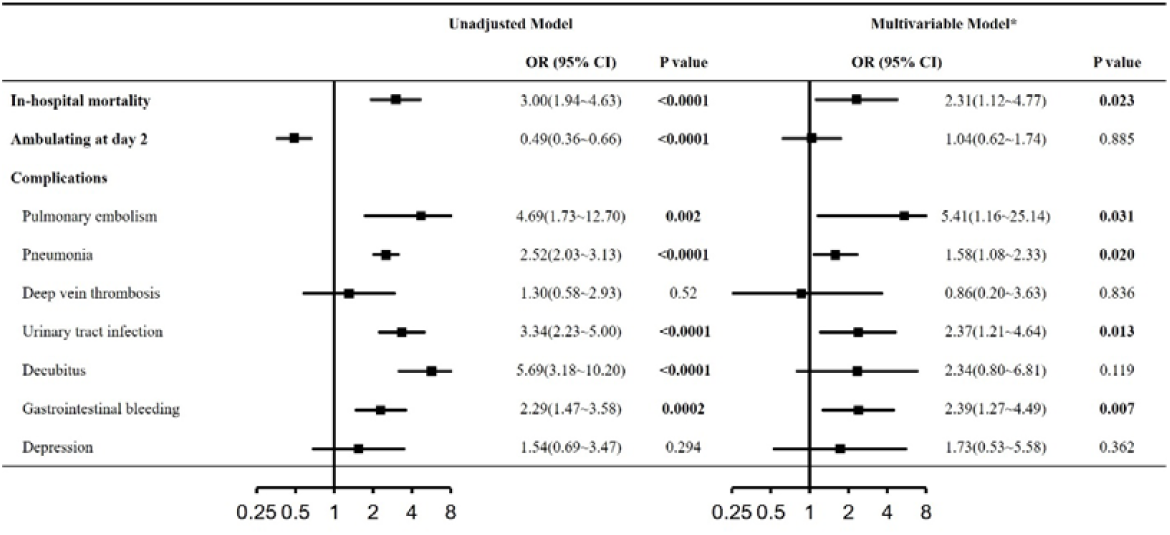
Logistic regression of the pre-stroke dementia on in-hospital mortality and discharge disposition

## Discussion

Findings from this study highlights the impact of pre-existing dementia on outcomes of patients with ICH and provides important quantitative estimates of such an association in a large and diverse Chinese population.

Limited data about preexisting cognitive status and stroke mortality are available. Besides preexisting cognitive status, low consciousness at stroke presentation, older age, and diabetes^6^ were reported to predict poorer ICH outcomes^6^. The Swedish Dementia Registry showed the preexisting dementia was associated with in-hospital stroke mortality^7^. And the association between pre-existing dementia with poorer outcomes (mortality) is significantly higher in Swedish than that in Chinese^7^. It may due to several reasons. Firstly, patient population recruited in the Swedish Dementia Registry were both ischemic and hemorrhagic strokes, while we recruited hemorrhagic stroke only. Secondly, the age was different between patient sample in Swedish Dementia Registry and the present study (age:80s vs 70s). The older age tended to have more severe strokes and more comorbidities in the Swedish group may explain severer intrahospital mortality. Both pre-stroke mild cognitive impairment and dementia were reported to be associated with increased mortality in stroke patients above the age of 65 years^6, 8^. Other three studies investigate the influence of pre-existing dementia on post-stroke mortality at 3 months or 1 year^4, 7, 9^. All these data indicated that pre-existing dementia was associated with 90-day case-fatality. Another study showed that Caucasians stroke patients (≤ 80 years) with preexisting Alzheimer’s disease or mixed dementia had higher mortality rates at 1 year compared to patients with preexisting vascular dementia^2^. Data about the relationship between preexisting dementia and long-term functional outcomes in Chinese lacked, further study should strengthen risk-factor management among patients with pre-existing dementia for stroke prevention.

In the current study, ICH patients with pre-existing dementia were observed to be older and had a greater frequency of multimorbidity which could indicate increased frailty and a greater propensity for poorer disease outcomes. Furthermore, ICH patients with pre-existing dementia could harbor concomitant neurodegenerative diseases such as Alzheimer’s disease and cerebral amyloid angiopathy, each capable of altering the brain cognitive connectomes and become more susceptible to dysregulation of the cerebrovascular dysregulation, decreased neuroplasticity, and cerebral ischemia^10^. All these pathologies would hinder recovery and increase the risk of mortality^11-13^. In addition, dementia could decrease participation of ICH patients in rehabilitation^13^ and their ability to recover lost functions and impaired neurogenesis^14^. Data from the current study suggests that patients with pre-existing dementia hade more in-hospital complications. These findings appear to differ from those in another study^15^ which showed no differences, probably due to the inclusion of a more diverse stroke population (hemorrhagic and ischemic stroke) compared to ours that comprised only those with hemorrhagic stroke^15^.

Our study has several limitations. First, data included the CSCA registry were mostly derived from larger tertiary centers with adequate resources to collect clinical information which may not necessarily reflect the broader context in rural hospital centers that may be less resourced. Second, although the diagnosis of dementia was made by a physician, information on dementia was based on self-reporting by patients and family members and could not be verified in all patients against medical records. Third, we were not able to identify the subtype of dementia (Alzheimer’s disease, vascular cognitive impairment, or mixed components). Fourth, neuroimaging information is not available from these patients, limiting our ability to correlate neuroimaging variables such as ICH volume and location and stroke outcomes. Future studies are necessary to elucidate the relationship between dementia and outcomes in ICH patients and evaluate the value of intensive risk factor control among individuals with pre-existing dementia for stroke prevention.

## Data Availability

The datasets during and/or analysed during the current study available from the corresponding author on reasonable request.

## Acknowledgements

The authors would like to thank all participants for their involvement.

## Author contributions

Lijun Zuo, Yang Hu and Yanhong Dong: study concept and design, analysis and interpretation of data, drafting of the manuscript. Lijun Zuo and Yang Hu served as the equally contributing first authors of the manuscript. and Hongqiu Gu: acquisition of data, analysis and interpretation of data. Raymond CS Seet, Zixiao Li and Yongjun Wang: acquisition of data and revision of the manuscript. Xingquan Zhao: obtaining funding, study concept and design, study supervision or coordination, revision of the drafting of the manuscript. All authors read and approved the final manuscript.

